# Characterization and Phase 1 Trial of a B Cell Activating Anti-CD73 Antibody for the Immunotherapy of COVID-19

**DOI:** 10.1101/2020.09.10.20191486

**Authors:** Stephen B. Willingham, Gerard Criner, Craig Hill, Shenshen Hu, Jenny A. Rudnick, Barbara Daine-Matsuoka, Jessica Hsieh, Haider Mashhedi, Andrew N. Hotson, Joshua Brody, Thomas Marron, Emily Piccione, Joseph J. Buggy, Suresh Mahabhashyam, William B. Jones, Mehrdad Mobasher, Richard A. Miller

**Author notes:** Corresponding author: Stephen Willingham, Corvus Pharmaceuticals.

## Abstract

COVID-19 is a global pandemic that has resulted in over 800,000 deaths. Robust humoral anti-viral immune responses have the potential to generate a diverse set of neutralizing antibodies to eliminate viruses and protect against re-infection, transmission, and the evolution of mutations that escape targeted therapeutics. CD73 is present on the majority of human B cells and a subset of T cells where it plays a role in lymphocyte activation and migration. CD73 also functions as an ectoenzyme that converts AMP into adenosine, which can be immunosuppressive. Here we report on CPI-006, a humanized FcγR binding-deficient IgG1 anti-CD73 antibody that blocks CD73 enzymatic activity and directly activates CD73^POS^ B cells, inducing differentiation into plasmablasts, immunoglobulin class switching, and antibody secretion independent of adenosine. Immunophenotypic analysis of peripheral blood from advanced cancer patients receiving CPI-006 revealed evidence of B cell activation, clonal expansion, and development of memory B cells. These immune effects suggested that CPI-006 may be effective at enhancing the magnitude, diversity, and duration of humoral and cellular responses to viruses such as SARS-CoV-2. We have therefore initiated a Phase 1, single-dose, dose-escalation trial in hospitalized patients with mild to moderate COVID-19. The objectives of this trial are to evaluate the safety of CPI-006 in COVID-19 patients and to determine effects of CPI-006 on anti-SARS-CoV-2 antibody responses and the development of memory B cell and T cells. Ten patients have been enrolled in the trial receiving doses of 0.3 mg/kg or 1.0 mg/kg. All evaluable patients had low pre-treatment serum levels of anti-viral antibodies to the SARS-CoV-2 trimeric spike protein and its receptor binding domain, independent of the duration of their COVID-19 related symptoms prior to enrollment. Anti-viral antibody responses were induced 7 days after CPI-006 treatment and titers continued to rise past Day 56. Increases in the frequency of memory B cells and effector/memory T cells were observed 28 days after treatment. These preliminary results suggest that CPI-006 activates B cells and may enhance and prolong anti-SARS-CoV-2 antibody responses in patients with COVID-19. This approach may be useful for treating COVID-19 or as an adjuvant to enhance the efficacy of vaccines.

## INTRODUCTION

Since its emergence in Hubei province, China, in December of 2019, the Severe Acute Respiratory Syndrome coronavirus 2 (SARS-CoV-2) and the associated coronavirus disease 2019 (COVID-19) has become a global health crisis.[1-3] There is an urgent need for therapies that can improve survival, clinical outcomes, and reduce the requirements for intensive supportive care and prolonged hospitalization.[4] A growing understanding of the viral/host interactions required for viral entry and replication has also informed the development of multiple vaccines.[5] The SARS-CoV-2 trimeric spike protein (TS) is a viral envelop glycoprotein which contains a receptor binding domain (RBD) in the spike 1 (S1) subunit.[6] This RBD directly interacts with N-terminal domain of angiotensin converting enzyme 2 (ACE2) on host cells to initiate a sequence of events leading to efficient viral infection.[3, 7] Multiple recent publications have demonstrated that antibodies that disrupt this interaction are effective at preventing viral infection.[6, 8] This provides evidence that neutralizing antibody responses to SARS-CoV-2 are possible and may provide clinical benefit in patients with COVID-19 and protection from disease in healthy subjects. Indeed, the U.S. Food and Drug Administration (FDA) has given emergency use authorization for the use of COVID-19 Convalescent Plasma (CCP) for the treatment of COVID-19. Clinical studies with CCP suggest that higher titers of neutralizing antibody provide superior clinical benefit to recipients.[9, 10] These findings support the value of anti-viral antibodies in eradicating viral infection in patients, lessening disease severity, and potentially reducing transmission.

CD73 is an emerging cancer immunotherapy target that was initially found to be important for lymphocyte trafficking and T cell activation.[11, 12] CD73 is expressed on subsets of human CD4^POS^ and CD8^POS^ T cells, germinal center follicular dendritic cells, and both naïve and class switched memory B cells.[12-14] A role for CD73 in B cell maturation has been proposed as reduced CD73 expression on B cells from patients with common variable immunodeficiency (CVID) correlates with an inability to produce IgG. [15, 16] Like many glycosyl phosphatidylinositol (GPI)-anchored molecules, CD73 has been shown to transmit activation signals when ligated by antibodies, although a physiologic ligand for CD73 has not been identified.[11, 12] CD73 also functions as an ectonucleotidase that hydrolyzes adenosine monophosphate (AMP) into adenosine.[17]

Here, we describe the generation and characterization of CPI-006, an IgG1k humanized FcγR binding-deficient anti-CD73 monoclonal antibody (mAb) that activates CD73^POS^ B cells. In vitro, CPI-006 induces the increased expression of markers associated with B cell maturation and antigen presentation, morphologic transformation to plasmablasts, and increased secretion of IgM and IgG. Biomarker studies in an ongoing Phase 1 cancer clinical trial revealed that CPI-006 causes a rapid and transient redistribution of circulating B cells that return to circulation enriched in a memory B cell phenotype.[18] Molecular studies of the B cell receptor (BCR) repertoire in treated patients suggests that CPI-006 stimulates the generation and expansion of novel B cell clones. These findings provided the rationale to examine CPI-006 as an immunotherapy for COVID-19 with the aim of enhancing anti-viral immune responses. The results of this trial to date suggest that the immunomodulatory properties of CPI-006 may be applied for the treatment of COVID-19 and also potentially utilized in combination with vaccines to induce immunity in healthy subjects.

## RESULTS

### CPI-006 Inhibits CD73 Enzyme Function and Reverses Adenosine-Mediate Immunosuppression

CPI-006 is a humanized anti-CD73 IgG1k antibody engineered with an N297Q mutation in CH2 of the heavy chain to eliminate Fc effector functions such as the ability to fix complement and initiate antibody dependent cellular cytotoxicity (ADCC, **Supplemental Figure 1A**). CPI-006 binds human CD73 with a K_D_ of 200 pM measured using bio-layer interferometry (**Supplemental Figure 1B**). The ability of this antibody to inhibit the enzymatic activity of cellular CD73 was evaluated by directly measuring free phosphate levels generated upon conversion of AMP to adenosine. CPI-006 inhibited CD73 activity to baseline levels that were established using the CRISPR CD73 knockout cell line and comparison to treatment with saturating amounts of APCP (adenosine 5'-(α,β-methylene) diphosphate), a small molecule inhibitor of CD73 enzymatic activity (**Figure 1A**). In contrast, MEDI9447, another anti-CD73 antibody that binds a non-overlapping epitope, only partially reduced CD73 activity.[19] CPI-006 also eliminated the enzymatic activity of CD73 expressed on primary human peripheral blood mononuclear cells (PBMCs) while MEDI9447 demonstrated a partial effect on enzymatic activity (**Figure 1B**). Collectively, these data show that CPI-006 and MEDI9447 inhibit catalytic activity of cellular CD73 expressed by tumor cells and lymphocytes, but only CPI-006 completely abolishes CD73 enzymatic activity.

**Figure 1:**
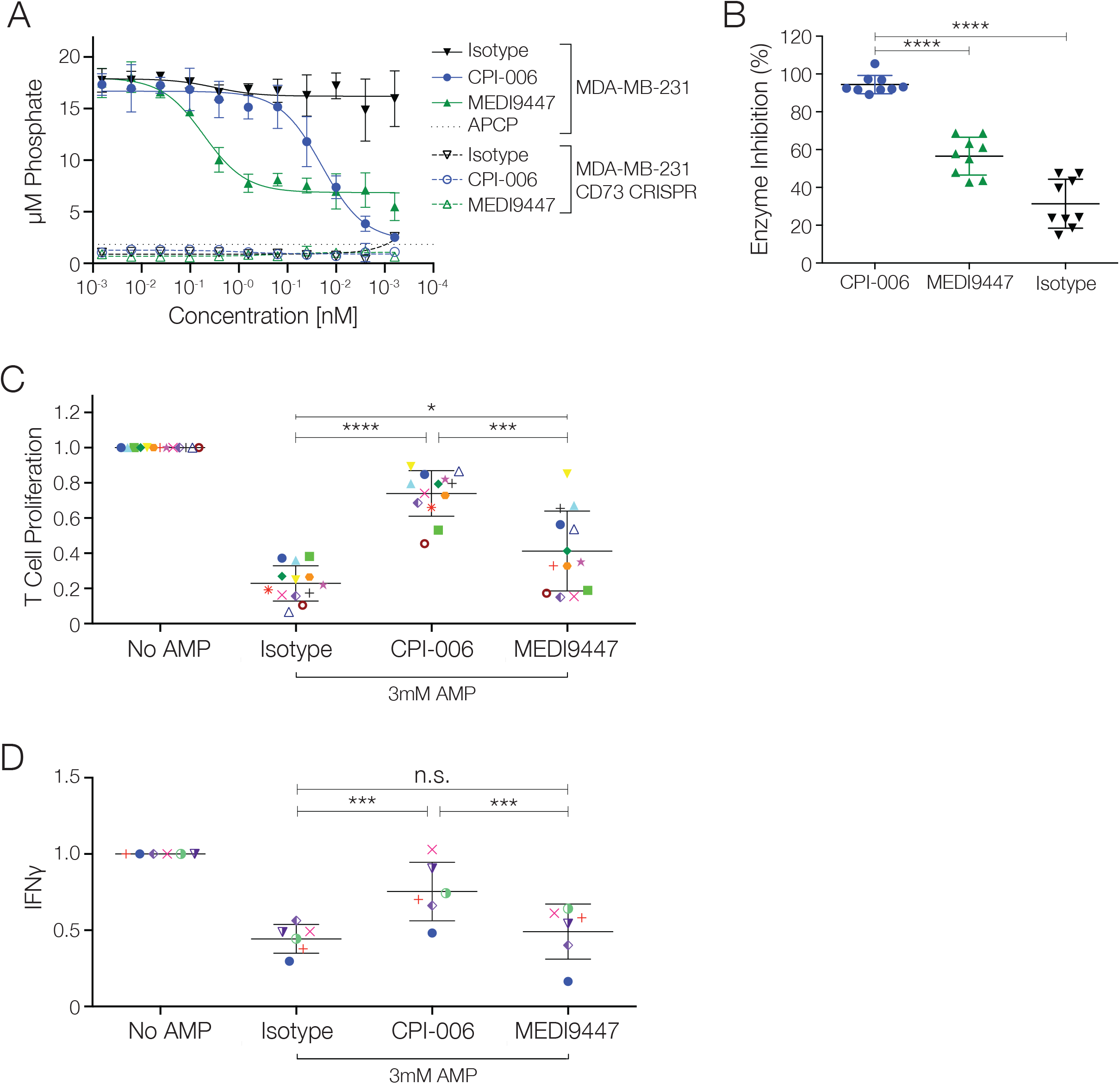
CPI-006 blocks CD73 enzymatic activity and restores T cell proliferation and cytokine production in an AMP-mediated immunosuppressive environment. A) CD73 catalytic activity was measured with MDA-MB-231 cells or MDA-MB-231 CD73 CRISPR cells in the presence of CPI-006, MEDI9447, or 1 mM APCP by adding 250 pM AMP and measuring phosphate levels in the cell culture supernatant. B) Human PBMCs were incubated with AMP in the presence of CPI-006, MEDI9447, or isotype control antibody at the saturating concentration of 300 nM. After 4 hours of incubation, the AMP remaining in the supernatant was quantified using the CellTiterGlo assay as described in the Methods. Percentage of inhibition was plotted. Data represent 9 individual donors and error bars represent mean ± SD. ****p<0.0001 as determined by t-test. C) Human PBMCs were activated with anti-CD3/anti-CD28 in the presence of AMP and a range of doses of CPI-006, MEDI9447, or isotype control antibody. T cell proliferation was assessed by flow cytometry. (C) T cell proliferation for 13 donors treated with 500 nM CPI-006 or MEDI9447 or 890 nM isotype control. Error bars represent mean ± SD. *p<0.05, ***p<0.001, and ****p<0.0001 as determined by t-test. (D) Human PBMCs were activated with anti-CD3/anti-CD28 in the presence of AMP and a range of doses of CPI-006, MEDI9447, or isotype control antibody. After 4 days in culture, IFNγ levels in the supernatant were measured by AlphaLISA (Perkin Elmer). IFNγ production for 6 donors treated as described in panel. Error bars represent mean ± SD. *p<0.05 and **p<0.01 as determined by t-test.

To model the immunosuppressive effects of adenosine on T cells, human PBMCs were obtained from healthy donors and cultured under T cell activating conditions. Addition of AMP resulted in decreased T cell proliferation and cytokine secretion. Addition of CPI-006 restored T cell proliferation in all donors tested, consistent with the antibody preventing conversion of AMP into immunosuppressive adenosine (**Figures 1C**). MEDI9447 restored T cell proliferation in a subset of donors but did not reach the magnitude of the CPI-006 response (**Figures 1C**). Interferon-gamma (IFNγ) production was evaluated as an additional readout for T cell response and similar results were observed (**Figures 1D**). These data demonstrate that antibody-mediated inhibition of CD73 activity blocks the suppressive effects of adenosine on T cell proliferation and cytokine secretion.

### CPI-006 Directly Activates Human B cells in vitro

CD73 is expressed on subsets of human hematopoietic cells and had previously been implicated to play a role in lymphocyte activation and adhesion [12, 13, 20, 21]. We performed a flow cytometry-based screen to identify differentially expressed cell surface markers on immune cells following in vitro treatment with CPI-006. CPI-006 strikingly activated B lymphocytes, resulting in the upregulation of activation markers (CD69 and CD83) and antigen presentation machinery (CD86 and MHC-II) to similar levels achieved with the positive control of BCR crosslinking via anti-IgM (**Figure 2A**). CPI-006 mediated B cell activation also resulted in increased cell surface expression of CD27, IgG, CD38, and CD138, all markers consistent with induction of B cell maturation (**Figure 2B**). CPI-006 induced activation was dose-dependent with concentrations of 1 μg/mL achieving near maximal induction of CD69 in vitro (**Figure 2C**). B cell activation has not previously been described in relation to CD73 signaling and is seemingly unique to CPI-006, as both MEDI9447 and clone AD2, two anti-CD73 antibodies that do not cross-block CPI-006, fail to induce activation (**Figure 2C**). Naïve B cells stimulated with CPI-006 in vitro underwent morphological changes consistent with differentiation into plasmablasts (**Figure 2D**). CPI-006 mediated B cell activation was restricted to CD73^POS^ B cells and required bivalent binding as CPI-006 immunoglobulin Fab fragments had minimal effect on expression levels of CD69 (**Figure 2E**). CPI-006 induction of CD69 was blocked with ibrutinib, a covalent BTK inhibitor (**Figure 2F**), demonstrating that CPI-006 directly activates B lymphocytes by invoking canonical B cell signaling pathways. B cell activation was not a consequence of the concentration of extracellular adenosine as addition of NECA (5'-(N-Ethylcarboxamido) adenosine), a potent and stable analogue of adenosine, did not block the induction of activation markers by CPI-006 (**Figure 2G**).[22]

**Figure 2:**
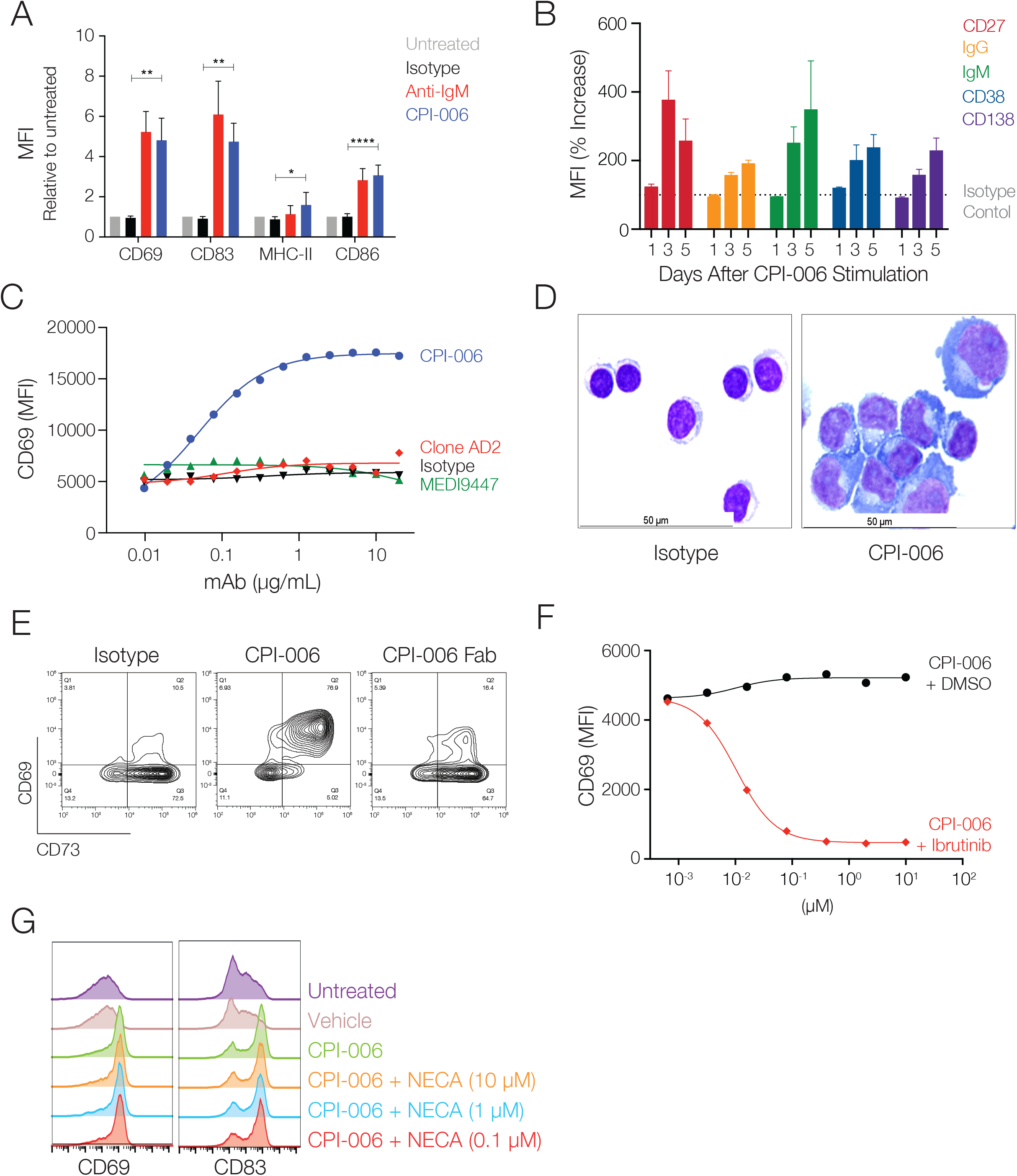
CPI-006 directly activates B lymphocytes and induces maturation into antibody secreting plasmablasts in vitro. (A) Purified B cells from 3-5 healthy donors were incubated overnight with human IgG1 isotype control or CPI-006 (10 μg/mL) or anti-IgM microbeads, a positive control for BCR stimulation. Expression of activation markers CD69, CD83, CD86, or MHC-II was measured by flow cytometry. B) Time dependent increases in the expression of CD27, IgG, IgM, CD38, and CD138 on purified B cells cultured in the presence of CPI-006 or isotype control (1 μg/mL). Mean fluorescence intensity (MFI) was determined for each marker and was normalized to the untreated or isotype control for each donor. C) B cell activation is unique to CPI-006 as other anti-CD73 antibodies do not induce CD69 expression. Expression of CD69 (MFI) was measured by flow cytometry. D) Representative images of purified B cells cultured with isotype control (left panel) or CPI-006 for 2 days. E) Purified B cells were incubated overnight with 10 μg/mL human IgG1 isotype control or CPI-006 or equimolar CPI-006 Fab. CD69 and CD73 were measured on B cells by flow cytometry. (F) Human PBMCs were incubated overnight with a fixed concentration of CPI-006 (10 μg/mL) along with ibrutinib or vehicle control over a range of concentrations. Expression of CD69 on B cells (CD19^POS^CD3^NEG^) was measured by flow cytometry. G) Human PBMCs were incubated overnight with 10 μg/mL CPI-006 with or without NECA over a range of concentrations or 10 μM APCP. Expression of CD69 on B cells (CD19+CD3-) was measured by flow cytometry and MFI is reported. Error bars represent mean ± SD. *p<0.05, **p<0.01 as determined by t-test.

To assess functional consequences of B cell activation with CPI-006, we first measured the concentration of IgG and IgM secreted by healthy donor PBMCs into the culture supernatant six days after in vitro treatment. Addition of CPI-006 resulted in a 3-fold increase in IgM and IgGλ levels (IgGκ not measured) relative to an isotype control, demonstrating that CPI-006 stimulates antibody secretion and possibly class switching to IgG (**Figure 3A**). CPI-006 also induced production of B cell cytokines such as CCL3, CCL4, CCL2, and CCL22 (**Figure 3B-3E**), but did not induce IFNγ, IL-2, IL-6, IL-10, or TNFα from healthy donor PBMCs (data not shown). CPI-006 would therefore not be expected to increase the potential for inflammatory cytokine release reported in some COVID-19 patients. [23-25] Collectively, these experiments demonstrate that CPI-006 activates B lymphocytes, resulting in morphological and immunological changes consistent with B cell differentiation and antibody production. Furthermore, this property is unique to CPI-006 and is independent of adenosine-modulatory activity.

**Figure 3:**
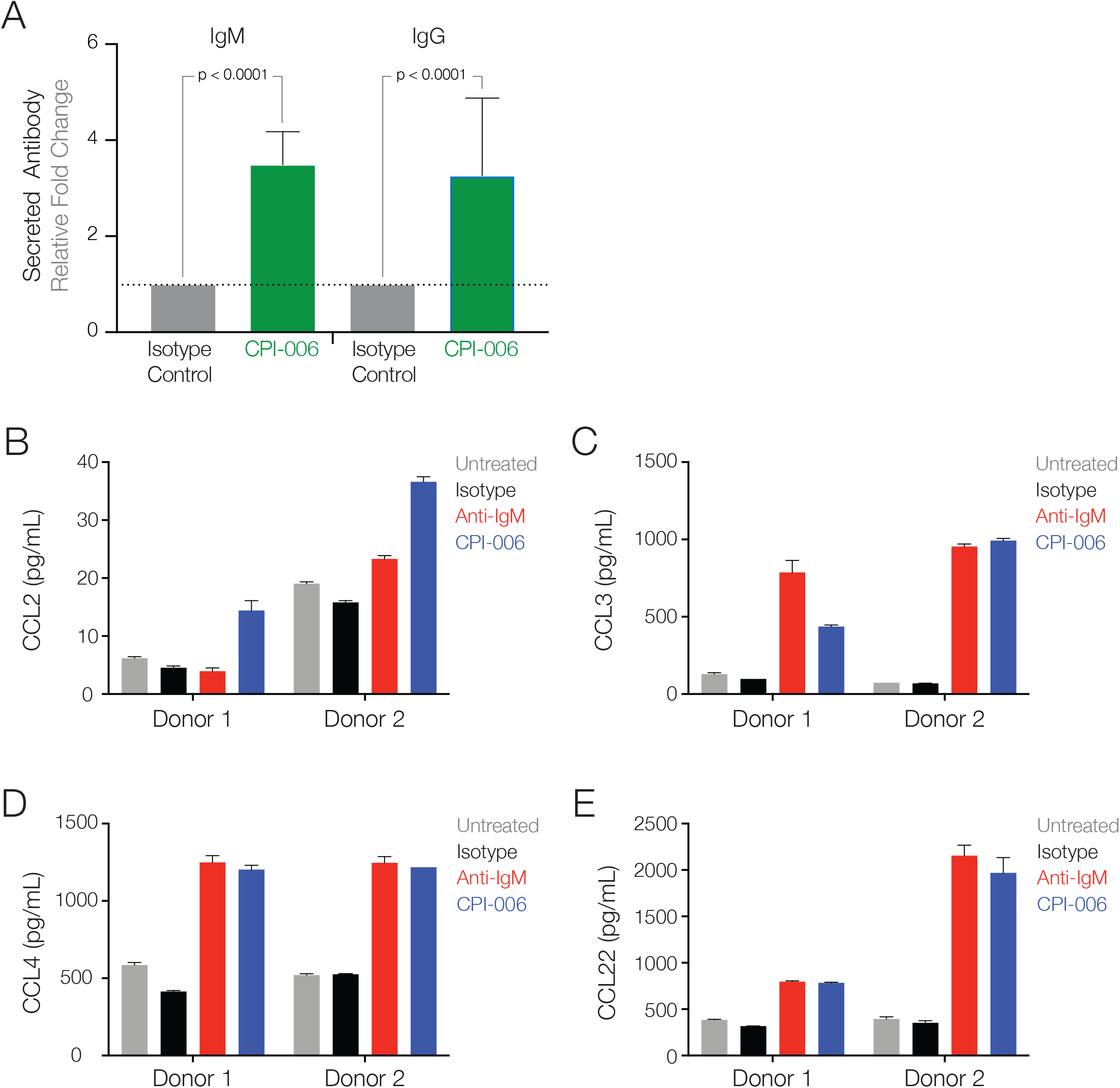
CPI-006 activates human B cells to secrete immunoglobulin and cytokines associated with B cell activation. A) Human PBMCs were incubated for 6 days with 1 μg/mL human IgG1 isotype control or CPI-006. IgGλ and IgM secreted into the culture supernatant was quantified by ELISA. IgGk was not measured as addition of CPI-006, an IgG1k antibody, would have confounded the results. Data are represented as mean ± SEM of 6 independent experiments, each in duplicate. B) Purified B cells were incubated for 5 days with 10 μg/mL human IgG1 isotype control or CPI-006 or anti-IgM microbeads, a positive control for BCR stimulation. Concentration of CCL2, CCL3, CCL4 and CCL22 in supernatant were measured by ELISA. Data are represented as mean ± SD of 4 independent experiments, each in duplicate.

### Immunologic Effects of CPI-006 in a Phase 1 Trial in Advanced Cancer Patients

CPI-006 is being evaluated as an immunotherapy for cancer in an ongoing phase 1 study (NCT03454451). In this dose escalation, repeat dose (21-day cycle) study, we observed dramatic decreases in circulating CD73^POS^ B cells at all CPI-006 dose levels (1-24 mg/kg), 30 minutes after antibody infusion (**Figure 4A**). CPI-006 does not induce B cell death or initiate ADCC (**Supplemental Figure 1A**) so this result is most likely due to the redistribution of activated B cells into lymphoid tissues. B cells returned to circulation at levels similar to baseline by day 21. Returning B cells were enriched in CD27^POS^ IgD^NEG^ class switched memory B cells in the majority of subjects receiving ≥ 3 mg/kg CPI-006 (**Figure 4B**). Memory B cells are essential for both acute and long-term immunity as they have undergone immunoglobulin rearrangement and somatic hypermutation in order to produce high affinity, antigen-specific antibody upon exposure to antigen. BCR repertoire analysis in a subset of patients revealed the emergence of novel B cell clones that were not present in the peripheral blood prior to treatment (**Figure 4C**). These new B cell clones were limited to 2-40 clones per patient and appeared with frequencies as high as 1:100 B cells, consistent with a robust clonal expansion following an antigen-specific response (**Figure 4C**). The cognate antigens recognized by these new B cell clones are currently under characterization. Representative CPI-006 treated patients (n=10) with robust diversification of BCR repertoire demonstrate unaltered antigen-specific IgG response against 5 common viruses (Mumps, CMV, Rubella, Measles, HSV1) at 21 days post treatment compared to baseline. This result demonstrates that CPI-006 does not simply induce polyclonal B cell activation and implies a requirement for concomitant antigen exposure in order for CPI-006 to induce an antigen-specific response (**Supplemental Figure 2**).

**Figure 4:**
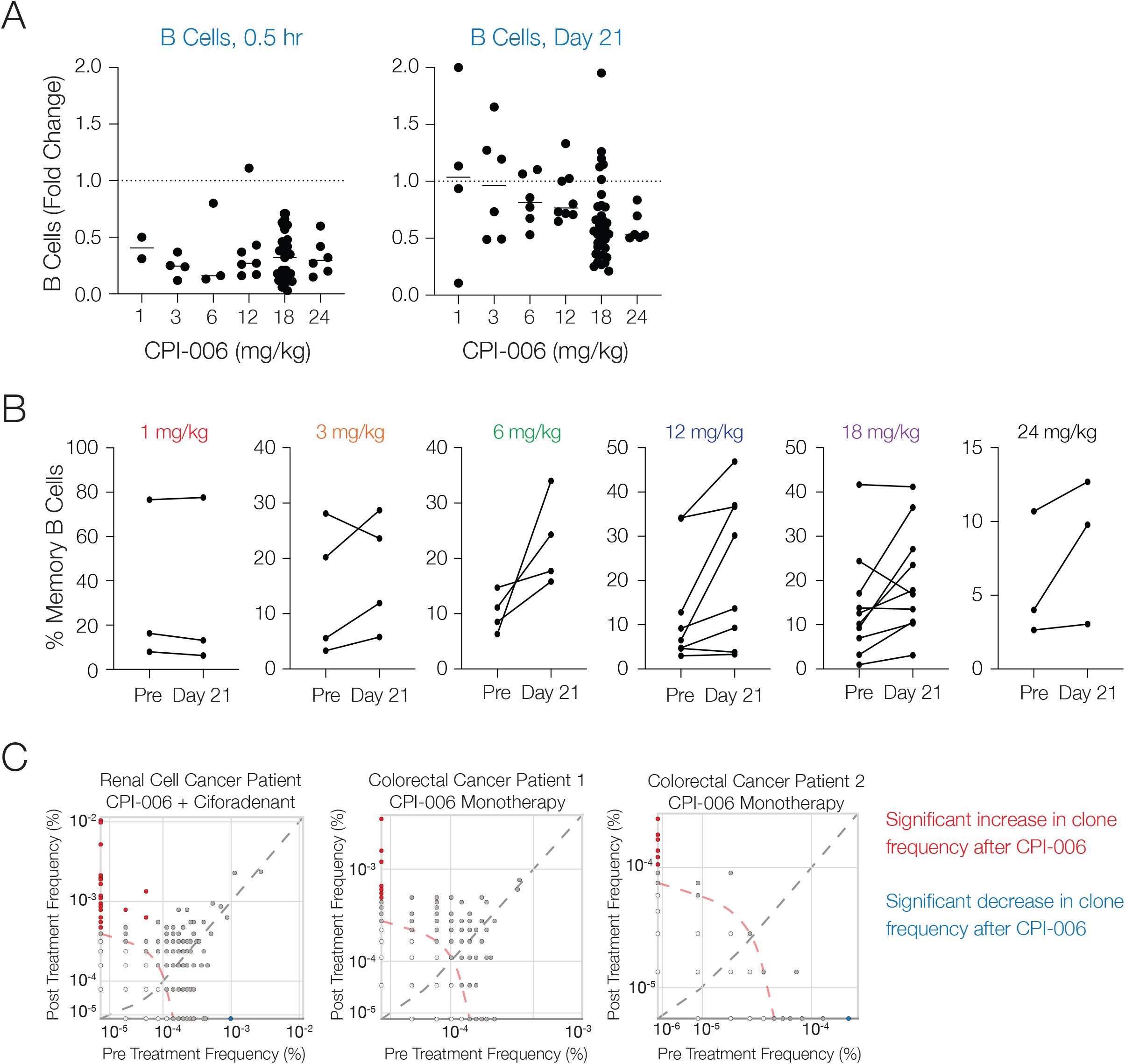
B and T cell dynamics in advanced cancer patients treated with CPI-006. A) CPI-006 induces rapid change in peripheral B cells, returning to baseline by Day 21. B) An increased frequency of memory B cells is observed at doses ≥3 mg/kg. Each symbol represents a treated patient, with lines connecting paired samples when appropriate. Some of these patients received an adenosine A2A receptor antagonist, ciforadenant, in combination with CPI-006. Studies with ciforadenant monotherapy reveal no changes in B cell numbers or immunophenotype. C) Representative examples of BCR repertoire diversification in 3 cancer patients 21 days after CPI-006 treatment.

### Immunotherapy of COVID-19 with CPI-006

#### Clinical Study Design

CPI-006-002 is a Phase 1 open label, dose-escalation trial evaluating the safety and immunologic effects of a single dose of CPI-006 as immunotherapy for hospitalized patients with mild to moderately severe COVID-19 (**NCT04464395**). SARS-CoV-2 infection is confirmed by RT-qPCR testing of nasal swabs and eligible patients must have blood oxygen saturation of at least 92% on ≤5L/min supplemental oxygen. Five patients per cohort receive doses of 0.3 mg/kg, 1.0 mg/kg, 3.0 mg/kg or 5.0 mg/kg delivered by intravenous infusion over 10-30 minutes. Patients are allowed to receive standard care for COVID-19, including remdesivir and steroids. Use of CCP or passively administered monoclonal anti-SARS-CoV-2 antibodies were not permitted. Safety and other disease assessments along with PBMC and serum collection are conducted at 7, 14, and 28 days, and at 2 months, 3 months and 6 months after receiving CPI-006.

#### Patient Characteristics and Safety

Five patients have been treated in the 0.3 mg/kg and five in the 1.0 mg/kg cohorts (Table 1). The median age was 64 years (range 28-76) and all had comorbidities including diabetes (4), hypertension (2), obesity (7,) and/or cancer (2). The median duration from presentation of symptoms (POS) to CPI-006 administration was 8 days (range 1-21 days). No infusion related reactions or other treatment related adverse events have been observed. All patients recovered with improvement of inflammatory markers and symptoms and were discharged at a median of 4 days after hospitalization (Table 1).

**Table 1:**
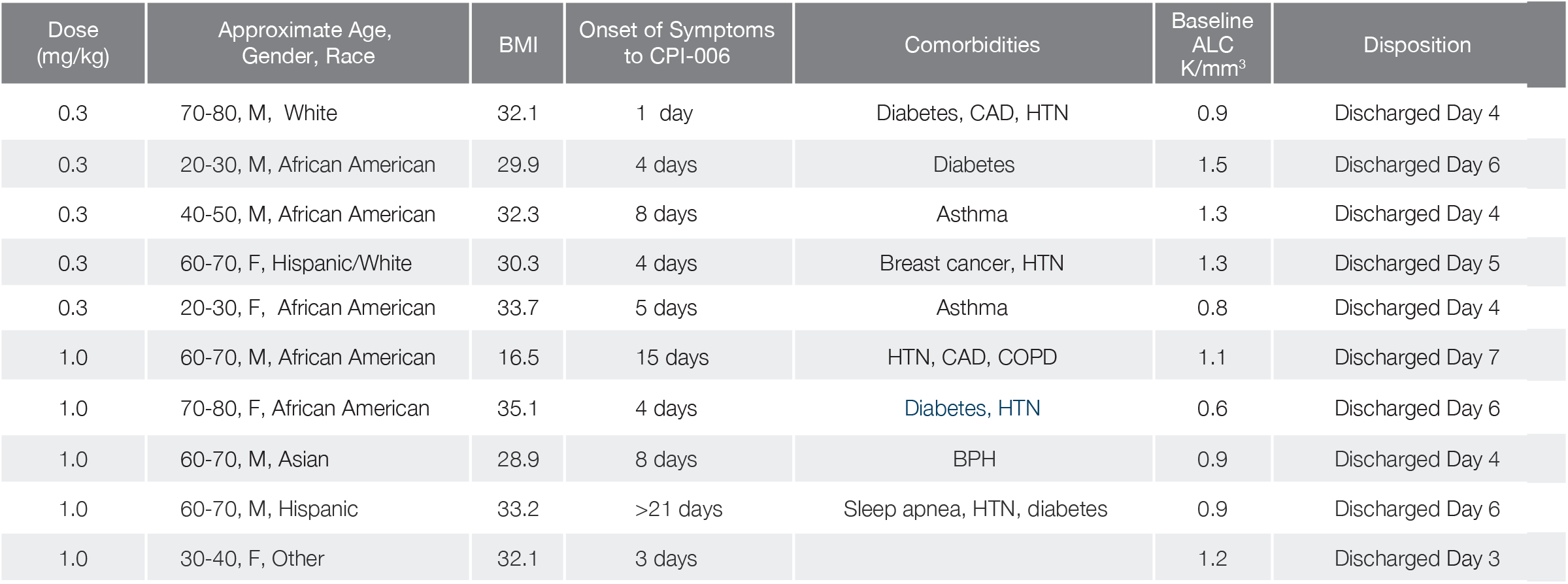
Baseline characteristics of enrolled COVID-19 patients treated with CPI-006. HTN - hypertension, CAD - coronary artery disease, COPD - chronic obstructive pulmonary disease, BPH - benign prostatic hyperplasia.

#### Immune Responses in CPI-006 Treated Patients

IgG and IgM antibody titers against the SARS-CoV-2 TS and/or RBD rapidly increased in 8/8 evaluable patients within 7 days of a single infusion of low doses of CPI-006 (**Figure 5A, 5B**). One patient did not have a pre-treatment sample available, but had a sample collected 24 hours after CPI-006 administration and this sample exhibited a high antibody titer. Similar results were observed in anti-SARS-Cov-2 IgA titers (data not shown). No correlation between time after POS and pre-treatment serum antibody levels has been observed as 9/9 evaluable patients had low pre-treatment titers despite relatively long durations of symptoms. For example, one patient had low titers at 21 days after onset of symptoms, but produced IgG and IgM titers rapidly rising to >100,000 and extending 28 days after administration of CPI-006, equivalent to 49 days POS. A corresponding increase in neutralizing antibody levels, measured by the ability to block recombinant RBD binding to human ACE2, was also observed (**Figure 5C**). Results in this assay have been shown by others to correlate with results in the live viral plaque reduction neutralization assay test (PRNTs).[26] Continually increasing IgG and IgM levels to viral TS or RBD antigens and neutralization antibodies were observed out to 28 days post treatment. It appears that prolonged high titer humoral response is possible as one patient has escalating levels out to 56 days. Antibody levels reached higher titers compared to convalescent sera from recovered patients with more favorable clinical characteristics (convalescent sera collected at 4-6 weeks POS, no comorbidities, and median age of 40) (**Figure 6A-6C**). Immunophenotyping of PBMCs at baseline and 14- or 28-days after treatment provided preliminary evidence that CPI-006 increased the frequency of memory B cells in two of two evaluated patients treated with 0.3 mg/kg (**Figure 7A**). An increased frequency of memory/effector CD4^POS^ and CD8^POS^ T cells was also observed (**Figure 7B-7C**) in three out of three evaluated patients. PBMCs stimulated with peptides derived from SARS-CoV-2 membrane (M), nucleocapsid (N), or spike (S) produced Th1 cytokines such as IFNγ and IL-2 following CPI-006 treatment in one patient evaluated thus far (**Figure 7D-7E**).

**Figure 5:**
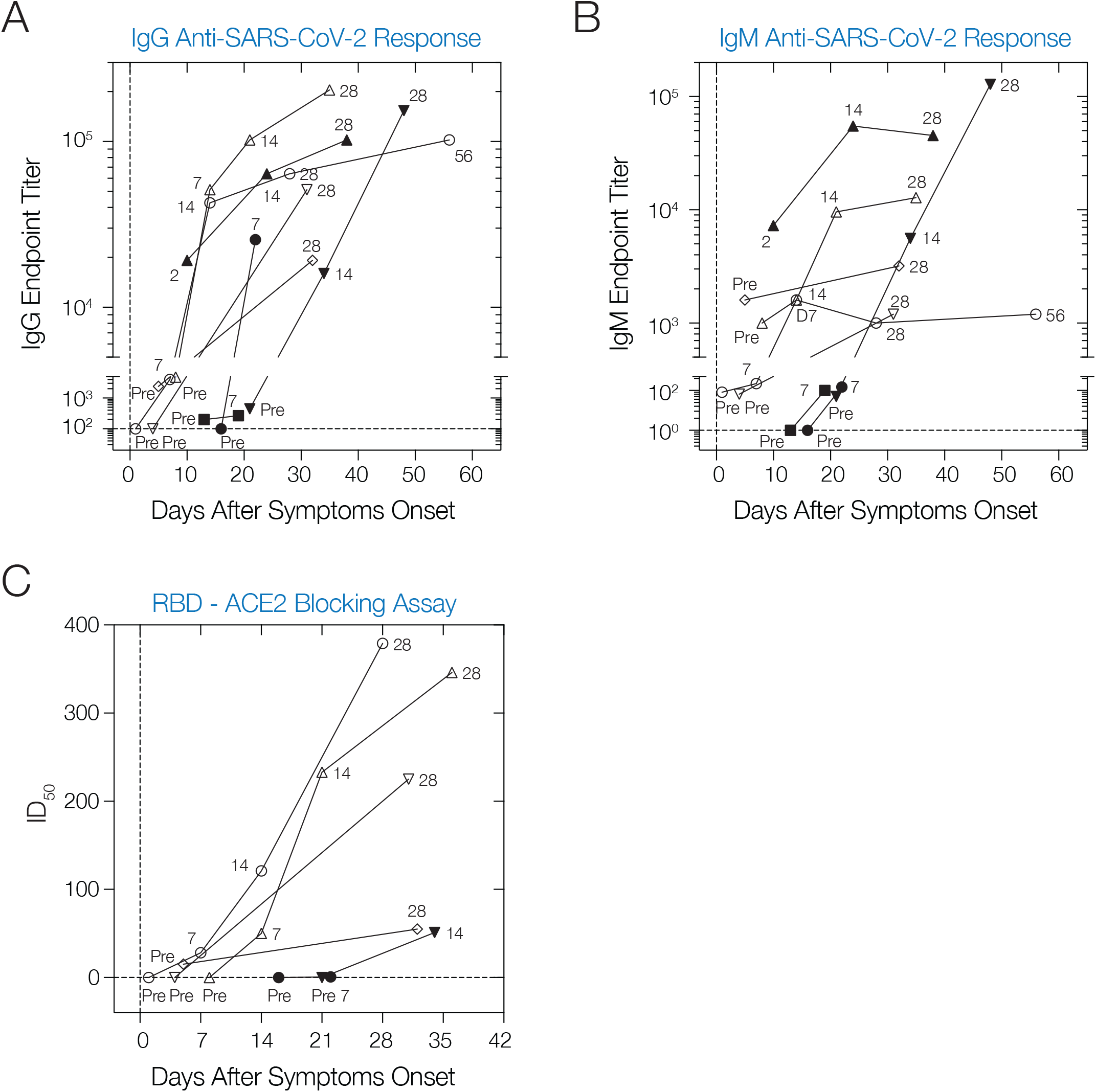
Anti-SARS-CoV-2 antibody responses in COVID-19 patients treated with CPI-006. A-B) Endpoint IgG (A) and IgM (B) titers directed to trimeric spike protein in eight COVID-19 patients treated with CPI-006. C) Inhibitory dilution required to block 50% (ID50) RBD binding to recombinant human ACE2. Each symbol represents an individual patient. Open symbols represent patients dosed with 0.3 mg/kg CPI-006. Filled symbols represent patients dosed with 1 mg/kg CPI-006.

**Figure 6:**
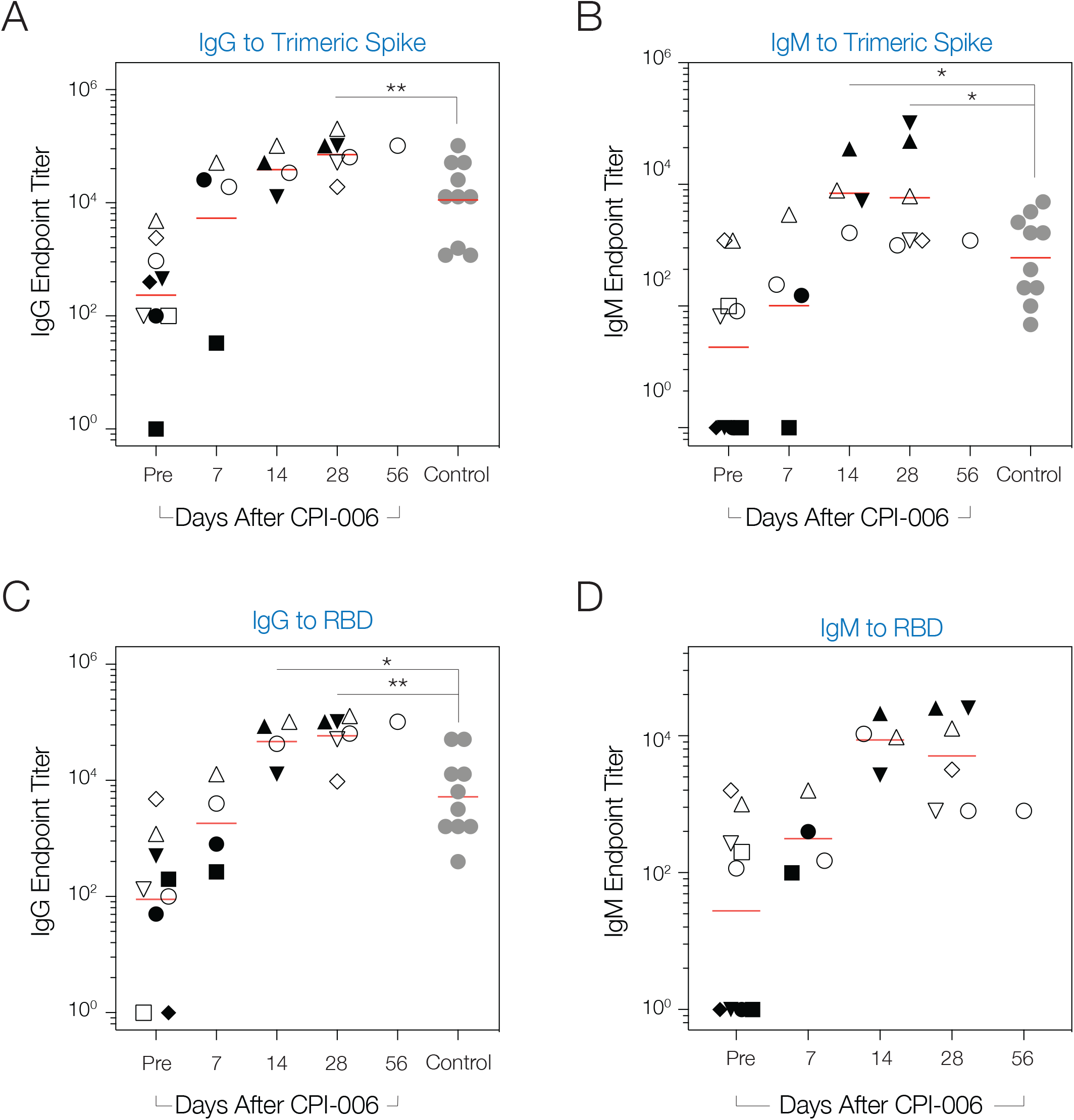
Endpoint titers of IgG and IgM directed to Spike and RBD in COVID-19 patients treated with CPI-006. A-B) IgG and IgM titers directed to recombinant trimeric spike protein. C-D) IgG and IgM titers directed to recombinant RBD. All titers were measured by ELISA. Each symbol represents an individual patient. Red line represents geometric mean of titers in each timepoint. Open symbols represent patients dosed with 0.3 mg/kg CPI-006. Filled black symbols represent patients dosed with 1 mg/kg CPI-006. Grey filled circles represent serum from untreated convalescent control individuals. *P* values between each timepoint and convalescent control group were determined with unpaired Welch's t-test of logarithm transformation of antibody titer values. All the statistical analyses were performed by GraphPad Prism 7. * p < 0.05; ** p <0.01.

**Figure 7:**
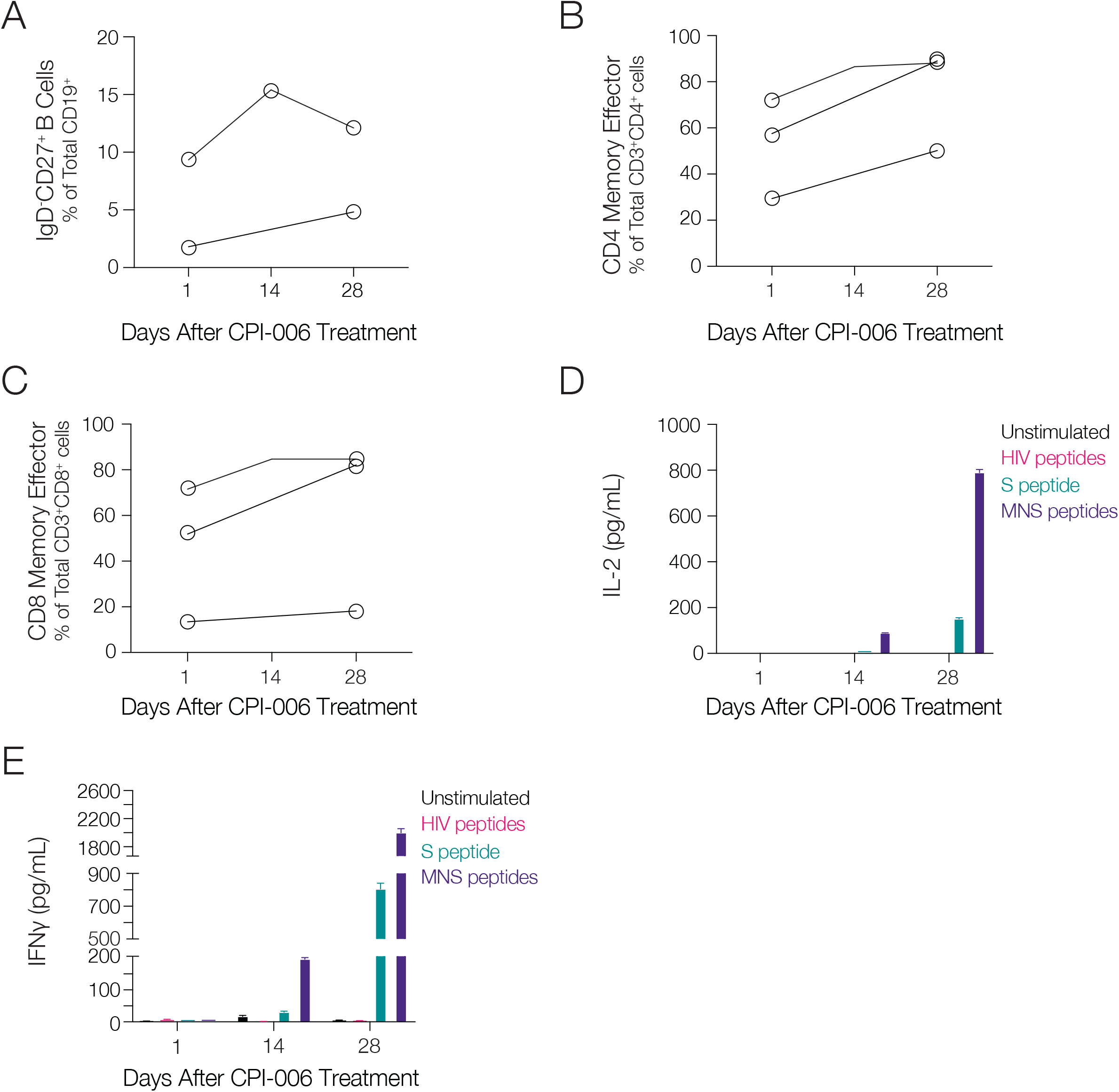
CPI-006 induces an increase in memory B cells and memory/effector T cells in COVID-19 patients. A) Frequency of memory B cells (CD19^POS^IgD^NEG^CD27^POS^) within CD19^POS^ gate at baseline and 28 days after treatment in two patients treated with 0.3 mg/kg CPI-006. A third patient treated with 0.3 mg/kg CPI-006 was also evaluated but had insufficient CD19^POS^ cells detectable at day 28 and was therefore excluded from the analysis. B-C) Increased frequency of memory/effector CD4^POS^ (B) and CD8^POS^ (C) T cells after CPI-006 treatment. Memory/effector population was defined as CD3^POS^CD45RA^NEG^. D-E) Serial evaluation of PBMCs from one patient treated with 0.3 mg/kg CPI-006 for ability to secrete IL-2 (D) and IFNγ (E) specifically in response to SARS-CoV-2 peptides but not control peptides derived from the HIV GAG protein. S peptide = Peptides derived from SARS-CoV-2 spike protein. MNS = Peptides derived from SARS-CoV-2 membrane, nucleocapsid, and spike proteins.

## DISCUSSION

CPI-006 is a humanized FcγR binding-deficient anti-CD73 mAb that directly activates CD73^POS^ B cells, thereby inducing their trafficking to lymphoid tissues and promoting antibody production and differentiation into memory B cells. CPI-006 induces markers of B cell activation (CD69), maturation (CD138, CD28), and antigen presentation (CD86, CD83, MHC-II) in vitro along with a corresponding morphologic transformation into antibody secreting plasmablasts. Studies in cancer patients demonstrated diversification of the BCR repertoire by stimulating the emergence and expansion of novel B cell clones. These findings indicate that CPI-006 can activate CD73^POS^ B cells to elicit antigen specific humoral and memory responses that display functional hallmarks associated with protective immunity. These effects are independent of CD73 enzymatic activity as addition of adenosine analogs or blockade of CD73 enzymatic activity alone has no effect on B cell activation in vitro. To our knowledge, CPI-006 is the only anti-CD73 antibody or small molecule inhibitor in development with the ability to directly activate B or T cells.

CD73 was originally characterized as a costimulatory molecule for T cells, but our results demonstrate that CPI-006 predominantly activates B cells.[11] Human CD73 is expressed on IgD^POS^IgM^DIM/NEG^ naïve B cells, and CD27^POS^ memory B cells expressing IgG or IgA.[14] CPI-006 induces the expression of CD69, an activation marker that negatively regulates S1PR1 function, resulting in the prolonged retention of activated B cells in lymphoid organs and thymus.[27] This increased lymphoid residence time provides time to complete B cell activation and interact with CD4^POS^ T follicular helper cells to shape downstream immune responses. While B cell activation with CPI-006 is independent of adenosine blockade in vitro, the CD73 enzymatic blockade may be complementary in vivo as adenosine has been shown to restrict lymphocyte migration into lymph nodes in preclinical animal models.[28] Additional studies will clarify if this temporary sequestration in lymph tissues is the dominant mechanism by which CPI-006 diversifies the BCR repertoire and promotes the development memory B and T cells.

This study demonstrates that antibody titers to TS and RBD with neutralizing activity toward RBD increase within 7 days in patients treated with a single, low dose of CPI-006. The single, low doses evaluated in our study thus far are noteworthy as the maximal concentrations of CPI-006 in plasma are modeled to exceed the 1 μg/ml threshold needed for maximal B cell activation yet low enough to be rapidly cleared. Although CPI-006 was delivered intravenously in this study, the effects seen with low doses indicate that alternative routes of delivery such as subcutaneous or intramuscular administration are feasible.

Most patients with COVID-19 become seropositive for IgG/IgM/IgA within 2-3 weeks following onset of symptoms.[29] However, all of the patients on this trial had low levels of anti-SARS-CoV2 antibodies at the time of hospitalization despite a wide range of duration of prior symptoms from 1 -21 days. The lack of response in these patients may be related to host factors reducing the ability to mount a humoral response or other unknown immunosuppressive effects of viral infection. CPI-006 may overcome this apparent immunodeficiency as we observed robust anti-SARS-CoV-2 antibody responses induced by CPI-006 in patients with long POS and low pre-treatment titers. The kinetics following seroconversion are still being clarified, but others have reported that antibody titers in COVID-19 patients plateau approximately 6 days after seroconversion before steadily declining in the following weeks (IgM/IgA) to months (IgG).[29] Our preliminary results show that anti-SARS-CoV2 titers continue to rise without plateauing in the weeks following treatment, possibly hinting at a more robust and durable humoral response that would theoretically improve clinical outcomes in COVID-19 patients. These effects may also serve to reduce viral transmission and expand the pool of qualified convalescent plasma donors.

Temporary protection against SARS-CoV-2 can be imparted by circulating neutralizing antibodies, but the key to long term immunity lies in the production of antigen-specific memory B and T cells capable of recognizing and eliminating any potential re-infection. Indeed, CD4^POS^ and CD8^POS^ T cells able to cross-react with SARS-CoV-2 were detected in patients 6 years after recovering from the original 2003 SARS-CoV outbreak.[30] The increased frequency in memory B cells we observed following CPI-006 treatment in cancer patients has thus far been measured in a very limited number of COVID-19 patients. Additional studies are underway to evaluate the antigen specificity of these memory B cells. An increase in the frequency of memory/effector T cells were also observed. Serial assessments of these populations will be required to determine the durability of immunological memory and whether a dose response emerges as we continue to escalate the CPI-006 dose. We note that imbalanced B and T cell responses have been reported in COVID-19 patients, including protective T cell responses without detectable seroconversion and, conversely, impaired CD4^POS^ T cell responses in critically ill patients with high IgG antibody titers. B cell recruitment and responses in peri-tumoral tertiary lymphoid structures has been associated with response to immunotherapy; thus, it's tempting to speculate that CPI-006 could help restore coordinated B and T cell responses in COVID-19 patients. [31-33]

Multiple viral and mRNA-based vaccines in development have demonstrated the potential to induce humoral and cellular responses to SARS-CoV-2 antigens in vaccinated normal subjects.[34, 35] While encouraging, rapidly diminishing antibody titers approximately two weeks after vaccination have necessitated a second booster vaccination in some instances.[34] CPI-006 is being developed as a therapeutic, but it is important to note that it could also be used as a vaccine adjuvant to boost the titer, diversity, and duration of antibody responses and potentiate the development of long term immunity by generating memory B and effector T cells. This combination approach may be useful to enable successful immunization with a single vaccination and particularly effective in vulnerable populations such as immunosuppressed and elderly patients who do not typically respond well to vaccines.

The use of CPI-006 in COVID-19 patients has several advantages over other therapeutic approaches. CPI-006 may stimulate enhanced production of anti-viral antibodies in SARS-CoV-2 infected patients to shorten both the disease duration and time to viral clearance. Such antibody responses are also likely to be polyclonal, generating antibodies reacting with multiple viral antigenic determinants, not only on TS but other viral proteins as well, that would minimize the probability of developing resistance through natural selection of epitope loss variants. The increase in memory B and T cells can also provide longer lasting immunity and prevention of re-infection and transmission. Other approaches such as anti-viral drugs or passively administered monoclonal antibodies can be compromised by the evolution of viral mutations able to escape therapeutic targeting. Monoclonal antibodies against SARS-CoV-2 will also require significant quantities of infused immunoglobulin(s) and may need repeated infusions in order to maintain concentrations required for passive immunity. In contrast, low doses of CPI-006 have the potential to be immediately deployed in order to induce a diverse repertoire of high titer anti-viral antibodies in patients exposed to SARS-CoV-2 or, potentially, other infectious disease. We note that diversifying the repertoire of antigen specific B cells would also facilitate the development of a broader cocktail of therapeutic mAbs when needed as these cells can be directly isolated, sequenced, and screened for antibodies targeting unique viral epitopes.[36]

Absent a randomized, double-blinded, placebo-controlled trial, we are currently unable to conclude that the effects observed thus far in treated COVID-19 patients are directly attributable to CPI-006. Our current results demonstrate an atypically robust and durable antibody response following CPI-006 treatment, but we cannot exclude the possibility that these responses would have developed naturally. Moreover, multiple confounding factors including the variability in patient responses, clinical setting, and lack of standardized testing methods complicate comparisons to other therapeutics in development. Nonetheless, these encouraging early results are in line with our hypothesized biological mechanism and we remain cautiously optimistic that longitudinal assessments of neutralizing antibody titers and B and T cell functional assays will clarify the contribution of CPI-006 to humoral and cellular immune responses. We encourage the reader to consider the results presented here as a work in progress that we intend to update regularly as additional safety, efficacy, and biomarker data become available.

## METHODS

See Supplemental Methods for additional details.

### Antibodies

CPI-006 was engineered by isolating VH and VL regions from the parental hybridoma generated by immunizing mice with human CD73 and screening for inhibition of CD73 activity. Humanization was performed by inserting CDRs isolated from the hybridoma into a human framework and the antibody was expressed as a human kappa/IgG1 antibody with the N297Q mutation introduced into the CH2 region to eliminate FcγR binding. MEDI9447 was cloned using the VH and VL chain sequences published in WO 2016/075099 AI application patent and was expressed as a human lambda/IgG1-TM antibody. Both antibodies were expressed in Expi-293 cells (Thermo Fisher Scientific) and purified by Protein A chromatography (HiTrap Protein A, GE Healthcare Life Sciences). For flow cytometry analysis, antibodies were labeled with AlexaFluor 647 (Life Technologies). Human IgG1 isotype control used for in vitro and vivo studies was purchased from Sigma-Aldrich and BioXCell, respectively. Fab fragments were prepared using the Pierce Fab Preparation kit (Thermo Fisher Scientific). Anti-CD73 antibody clone AD2 was purchased from Abcam.

### Affinity measurements

Binding experiments were performed on Octet HTX at 25°C. Purified antibodies (2 |j,g/mL) were loaded onto Anti-Human IgG Fc Capture biosensors. Loaded sensors were dipped into a threefold dilution series of antigen (starting at 300 nM). Kinetic constants were calculated using a monovalent (1:1) binding model.

### Anti-SARS-CoV-2 antibody ELISA assays

ELISA was performed to measure the IgG, IgM, IgA antibody titer to the receptor-binding domain (RBD) of the spike protein and full-length spike trimer of the SARS-CoV-2 virus. Purified recombinant SARS-CoV-2 RBD and full-length spike protein were obtained from LakePharma. Briefly, ELISA plates were coated with RBD or spike protein (2 μg/mL in PBS) overnight at 4°C then blocked with 3% BSA in PBS for 1 hr. at room temperature (RT) after three washes with PBST. Serial dilutions of serum were prepared in PBST containing 1% BSA, then dispensed to the wells of the coated microtiter plate and incubated for 2 hr at RT. After three washes, the bound antibody was detected using anti-human IgG-horseradish peroxidase (HRP) conjugated secondary antibody (1:3000, Sigma-Aldrich, A0170) or anti-human IgM HPR secondary antibody (1:3000, Sigma-Aldrich, A0420), or anti-human IgA HRP secondary antibody for 1 hr at RT. After three washes, the reaction was developed by the addition of the substrate o-phenylenediamine dihydrochloride (SigmaFast OPD) and stopped by HCl (2M). The absorbance at 490 nm (OD490) was measured using an Envision plate reader (PerkinElmer). Recovered COVID-19 patient serum was obtained from Sanguine Biosciences from recovered patients confirmed to have had a COVID-19 PCR+ result. All serum samples were treated to inactivate infectious virus by incubation at 56°C for 30 mins. Healthy volunteer serum samples obtained from Stanford Blood Center during 2018 served as negative control. The titer cutoff value at OD490 is the mean plus 3 standard deviations of the negative controls. The endpoint titer is reported as the highest dilution of at least two before the OD490 decreases below the cut-off value. Data were analyzed using GraphPad Prism 7.

### RBD-ACE2 blocking assay

A MaxiSORP ELISA plate (Nunc) was pre-coated with human ACE2 protein (GenScript) at 100 ng per well in 50 μL of 100 mM carbonate-bicarbonate coating buffer (pH 9.6) overnight at 4 °C, followed by blocking with OptEIA assay diluent (BD). Human sera were prepared using a 2.5-fold serial dilution starting at 1:5. The sera was diluted a further 2-fold by addition of an equal volume of HRP-conjugated RBD (Genscript) and the mixture incubated for 30 min at 37 °C in a final volume of 150 μL. The mixture (100 μL) was transferred to the wells of the precoated ELISA plate and incubated for 15 min at 37 °C. Unbound HRP-RBD was removed by washing the plate four times with 260 μL of phosphate-buffered saline, 0.05% Tween-20. Bound antigen was detected by addition of 100 μL of the chromogenic substrate, 3,3',5,5'-tetramethylbenzidine followed by incubation for 20 min at 22 °C. The reaction was quenched by addition of 50 μL of 3 N HCl and the absorbance at 450 nm read using an EnVision plate reader (PerkinElmer). ID50 values were obtained by fitting the response-normalized data to a four-parameter logistic equation using GraphPad Prism version 8.4.3 for Windows, GraphPad Software, San Diego, California USA.

### Antigen specific T cell peptivator assay

Cryopreserved PBMCs from pre-2020 control donors or CPI-006 treated COVID-19 patients were thawed, washed, and incubated in RPMI 1640 (ATCC, Catalog #30-2001) plus 5% human serum (Sigma, Catalog #H4522-100ML) with Pen/Strep (Gibco, Catalog#15140122) for 24 hours with 600 nM SARS-CoV-2 peptides including S peptide (Miltenyi Biotec 130-126-700), M peptide (Miltenyi Biotec, Catalog #130-126-702), N peptide (Miltenyi Biotec, Catalog #130-126-698) or control HIV peptides (JPT, Catalog #PM-HIV-CONB). Culture supernatants were collected and cytokines were assayed according to manufacturer's protocol using the V-PLEX Proinflammatory Panel 1 Human Kit (MSD, Catalog #15049D)

### Immunophenotyping of CPI-006 treated patient PBMCs

PBMCs from COVID-19 patients were isolated within 24hr of blood collection and cryopreserved for flow cytometry. Expression of cell surface markers associated with B and T cell activation were assessed by flow cytometry using Fc blocking reagent (Miltenyi Biotech, Catalog #130-059-901) and antibodies directed to CD19 BV421 (Clone HIB19; BD Biosciences, Cat #562440), CD38 BV510 (Clone HB-7; BioLegend, Cat #356612), IgD FITC (Clone IA6-2; BD Biosciences, Cat #555778), CD73 PE (Clone AD2; BD Biosciences, Cat #550257), Mouse anti-human PD-1 PerCP 5.5 (clone EH12.1, BD Biosciences, Catalog #561273), CD3 PE-Cy7 (Clone UCHT1; BioLegend, Cat #300420), CD27 APC (Clone L128; BD Biosciences, Cat #337169), Mouse anti-human CD45RA APC-Fire 750 (clone HI100, BioLegend, Catalog #304152), Mouse anti-human CD8 BVD650 (clone SK1, BD Biosciences, Catalog #565289), Mouse anti-human CD4 PE-CF594 (clone SK3, BD Biosciences, Catalog #566914), Rat anti-human CXCR5 APC-R700 (clone RF8B2, BD Biosciences, Catalog #565191). Cryopreserved PBMCs from COVID-19 patients were analyzed at Corvus Pharmaceuticals using a CytoFLEX (Beckman Coulter). Fresh blood from cancer patients were analyzed at ICON using the same antibody clones. Flow data was analyzed using FlowJo v10.7.

### B cell receptor analysis

Sequencing of the CDR3 regions of human variable chains was performed using the immunoSEQ® BCR Assay (Adaptive Biotechnologies, Seattle, WA). Genomic DNA was extracted from PBMCs and was amplified in a bias-controlled multiplex PCR, followed by high-throughput sequencing and the abundance of each unique BCR region was quantified.[37-39]

## Data Availability

Further information and requests for resources and/or reagents should be directed to Dr. Stephen Willingham.

## ACKNOWLEDGMENTS

We thank Brandon Dezewiecki, Felica Flicker, Chunyan Gu, James Janc, Janet Koe, Long Kwei, Jennifer Law, Liang Liu, Gabriel Luciano, Sahil Malpotra, Brian Munneke, Florentino San Pablo, Deborah Strahs, Bradley Wolfe, Katherine Woodward, Jingrong Xu, and other current and former Corvus employees for administrative, clinical, and scientific support. We thank the patients, their families, and the physicians, nurses, and staff at Temple University Hospital and Mount Sinai Hospital for their participation in these clinical studies.

